# Covid-19 pandemic Analysis using Regression

**DOI:** 10.1101/2020.10.08.20208991

**Authors:** P Raji, GR Deeba Lakshmi

## Abstract

Covid-19 is a pandemic which has affected all parts of the world. Covid-19 is a pandemic which can be controlled only by maintaining social distancing, proper hygiene, wearing mask, hand sanitation and to a extend by wearing face shield. Even though each state has followed their own ways of controlling the infection, awareness among citizens and behaving as responsible citizens is very important in controlling this disease. Contact tracing plays an important role in controlling this pandemic. This paper deals with the effect of Covid-19 in various states of India and also forecasts its effect using machine learning techniques. Regression analysis like Linear and polynomial have been used for analysis of Covid-19, where Kaggle dataset has been used. This helps in understanding the much-affected states in India and helps to understand the infrastructure requirements to handle this pandemic efficiently.

## 1. Introduction

COVID -19 was initially originated in bats and later transmitted to humans. Also it can transmit between people. Other pandemic similar to COVID was had emerged in Guangdong, China in 2002 and emerged in Middle East in 2012 like SARS and MARS.SARS-CoV-2 emerged in Wuhan in 2019.Since the day of its outbreak it has affected more than 200 countries. Since the day of outbreak, there have been 15,83,792 confirmed cases and 34,968 deaths in India as on 31^st^ July 2020.

Machine learning helps computers to learn, without being programmed. Machine learning is classified to two types, supervised learning and unsupervised learning. In supervised learning, new examples are mapped by analysing input output relationship of the training data. Auto regression techniques, SEIR models and seasonal periodic regression model are used to predict Covid-19 outbreaks [1][2][3]. Suspected-Infected-Recovered-Dead (SIRD) Model [9] estimates epidemiological parameters like recovery and mortality rates, reproduction number and infection. Also, Long short-term memory and Gated Recurrent Unit using Recurrent Neural Network [15] has been used to predict recovered, negative cases, death cases and confirmed cases. In this paper, linear and polynomial regression models are created using number of confirmed cases, number of deaths and recovered cases. These models can help to predict the expected number of cases and deaths in the coming days.

Covid -19 is a new infection that’s been threat to the human life. Coronavirus are in the range of 80-160 nanometers size. Very basic signs of Covid-19 are high temperature and Breathing faster than usual. Symptoms of Covid-19 are Fatigue, Nausea, Loss of taste or smell, Muscle Ache, Cough, Headache, Sore Throat, Tiredness, chills and Fever. There are some cases where people don’t show any symptoms and are called asymptomatic. Very specific symptom of Covid-29 is the loss of ability to taste or smell which is very rare in other infections.

Patients should immediately seek medical care if there are warning signs or symptoms like

i. Blue lips or face, which could mean they are not getting enough oxygen.
ii. Chest pain when breathing
iii. Shortness of breath.
iv. New confusion or difficulty waking up.

People who are more affected by covid-19 are older adults greater than 65 years of age, obese and who have diabetes, hypertension or other ailments. So it’s better that these category people quarantine from other members in the family.

50% of people infected with SARS-Covid-2 will become ill within 5 days after they get infected. People who are asymptomatic can also be infectious. It’s more difficult to define infectious period for such people. The risk for death depends on access to care and general health. People who are in the age of 65 to 75 years old, almost 2% dies.4% to 10% dies who are at the age of 75 to 85 years. More than 10% dies if the age is above 85 years.

If infected by Covid-19, it can affect the lungs. Patients cannot breathe on their own if the lungs cannot recover. Lack of oxygen can damage the organs in the body causing increased risk of heart attacks, kidney failure, strokes and clotting disorders.

There are two types of tests used to identify Covid-19, Diagnostic test and Antibody test. Diagnostic tests are polymerase chain reaction (PCR) tests used to identity the presence of virus in the body. It detects the RNA of the virus. This is tested among people who shows sign and symptoms. Swab is taken from nose, throat and mouth. Also known as molecular tests helps to know whether the virus is reproducing in the cells. Antibody test identifies the presence of antibodies to the virus in the blood. The body starts to produce IgG antibodies 10 to 14 days after infection. This test is usually conducted on blood. This test is usually performed on people who have recovered or never had any symptoms.

Reproductive Number gives a measure of how fast the disease can spread. The higher the reproductive number, the more people will be infected. The R0 of SARS-COvid-19 is 2 to 3. If each infected person infects just two people, the size of outbreak doubles quickly. So contact tracing is also very important to reduce the transmission of Covid 19. Different countries are following different methods of electronic media to trace contacts. India is a country where citizen privacy is given utmost importance. There also many apps which are used to identify whether a person was exposed to Covid positive patient. But efficiency of this depends on the number of people using the app and also the data should be updated by all who are using it.

If the contacts can be traced efficiently and depending on the situation, if they can be isolated or quarantined, the spread of the disease can be controlled to a great extent.

## 2. Methods

Regression analysis has been used to understand the effect of Covid-19 on various Indian states. India is a country where Initially Covid-19 infection was controlled through lockdown of entire country and later by restricted lockdown. As the number of cases increased, depending on clusters lockdown is imposed.

**Figure 1:**
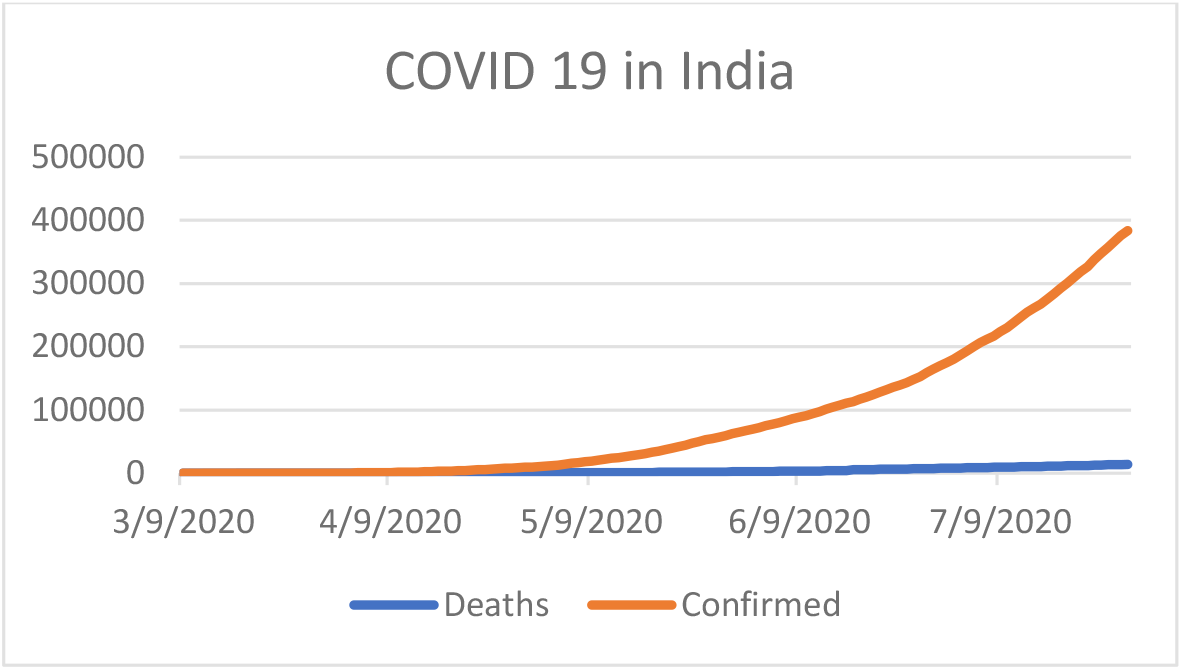
Confirmed and Recovered cases of Covid -19 of India.

Cases in various states of India has been analysed in terms of confirmed cases, death cases and recovered cases. Dataset has been taken from Kaggle website from 2^nd^ March 2020 to 28^th^ July 2020.

The number of cases in Tamil Nadu is much more compared to other four states and Kerala have been the least for the considered time period.

**Fig 2.**
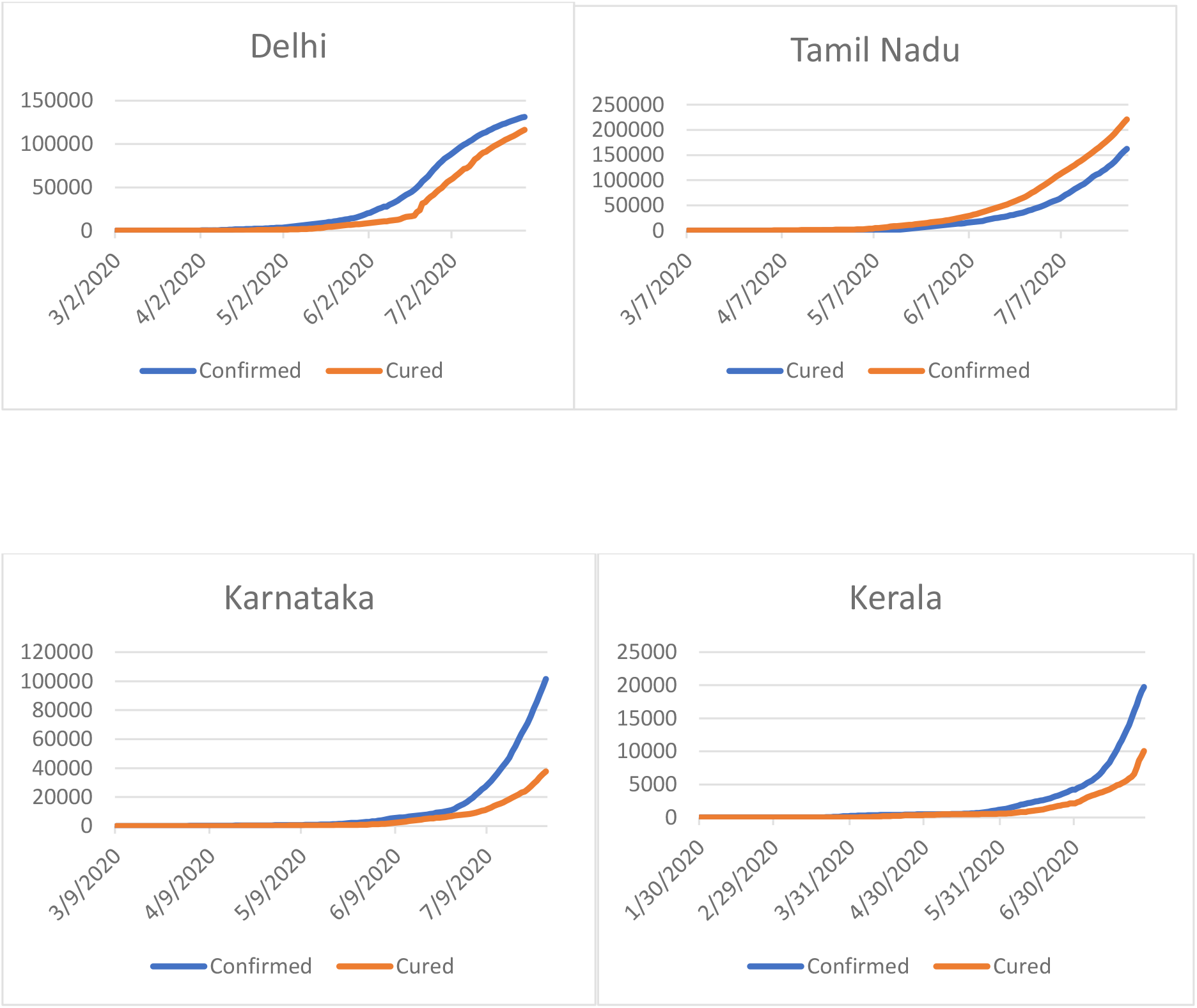
Covid-19 confirmed and cured cases in different states of India.

Kerala has been a state which received much appreciation world wide for initially flattening the curve.But later Kerala also lost his control over infection rate.The recovery rate of Delhi is much better compared to other states.There has been a steep increase in cases in all the states after the month of June.

Linear regression analysis and polynomial regression analysis has been modelled using Matlab software and effect of Covid-19 has been analysed.A relationship between variables are modelled using regression analysis.The variables considered are confirmed cases and mortality rates.

## 3. Results

The results of linear regression are shown in below figures.The results for karnataka almost matched with training set, but in case of other states there is slight deviations.Initially all the states tried to control the disease by quarantining the prople who got infected.But in case of all the states the number of cases are increasing.The mortality rate of kerala is much less compared to other states. Kerala had initiallycontrolled by very efficiently tracing the contacts.

**Figure 3:**
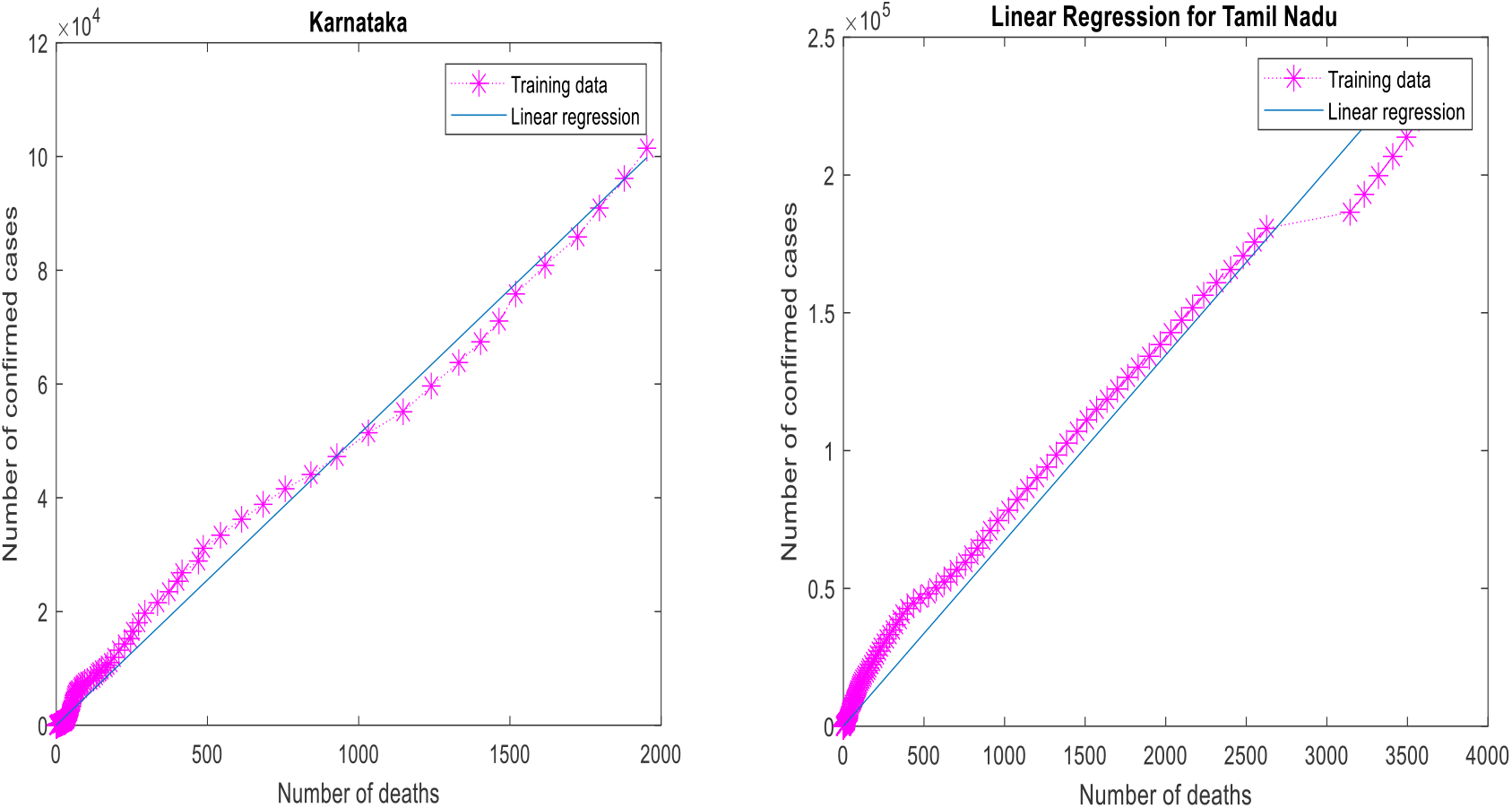
Linear Regression for Karnataka and Tamil Nadu.

**Figure 4:**
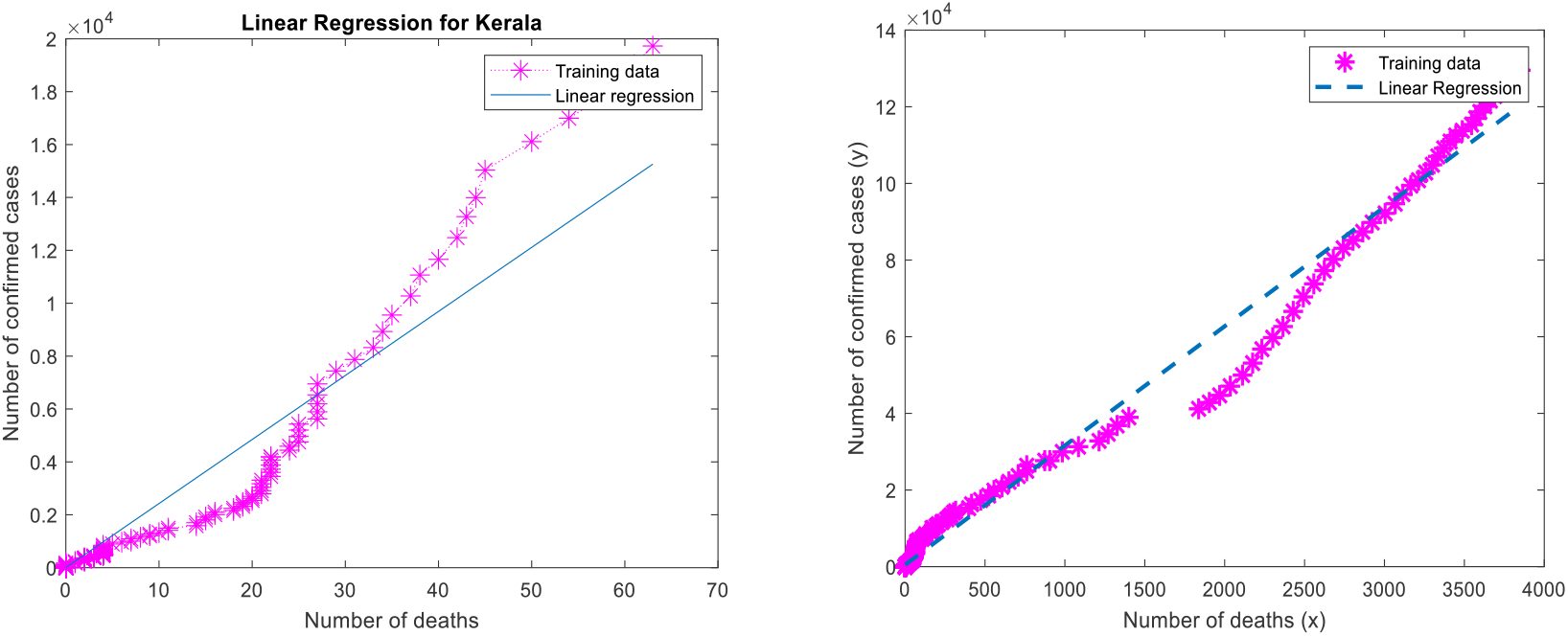
Linear regression fit for Kerala and Delhi.

In case of Karnataka, for above 12,000 cases there are chances of nearly 2000 deaths.The number of cases of Tamil Nadu is much more compared to other states.The mortality rate of Tamil Nadu and Delhi is almost same even though the number of cases in Tamil Nadu is much more than Delhi.

As on 3^rd^ August the number of confirmed cases in Karnataka is 134818 and deaths are 2496.In Delhi the number of confirmed is 137677 and deaths are 4004.In Tamil Nadu, 4132 has lost their lives and 257613 are confirmed cases. In Kerala, number of confirmed cases is 25911 and number of death is 82 which is much less compared to other states. Maharashtra has the highest number of cases and mortality rate.

**Figure 5:**
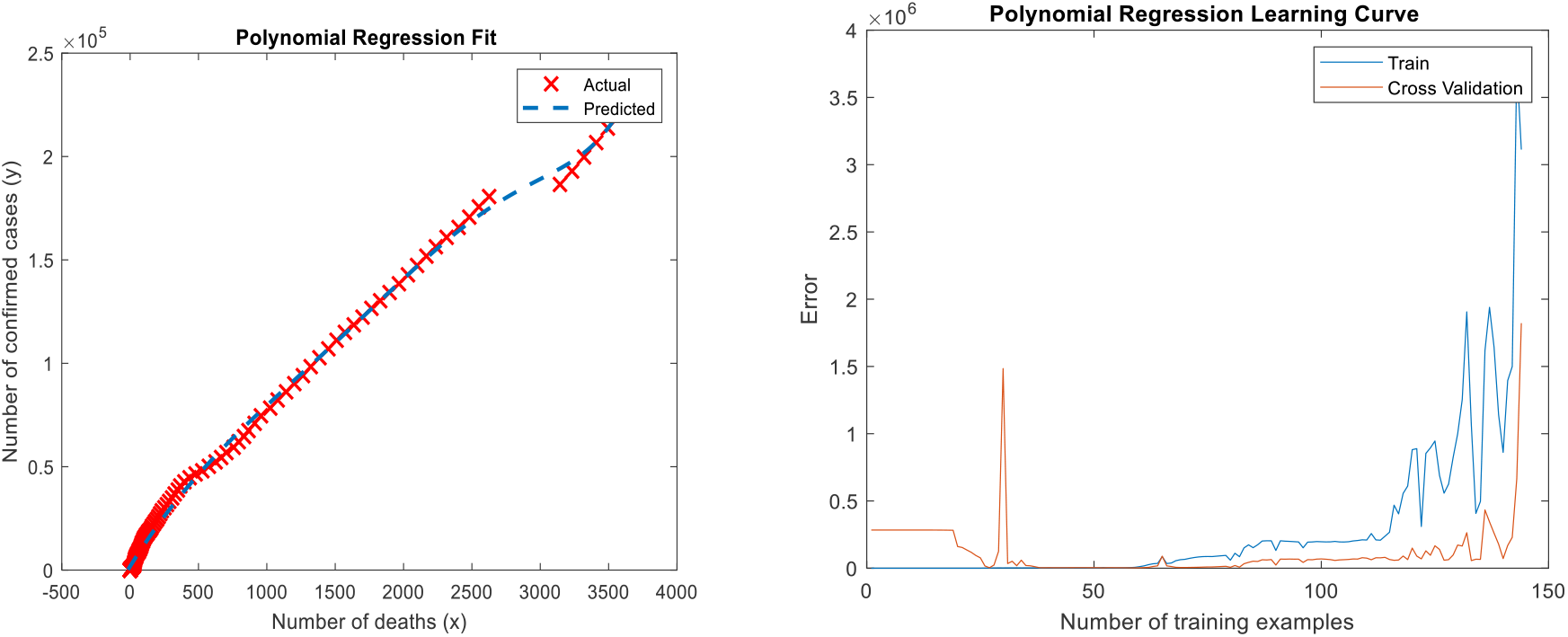
Polynomial Regression fit for state of Tamil Nadu and Polynomial Regression Learning Curve.

**Figure 6:**
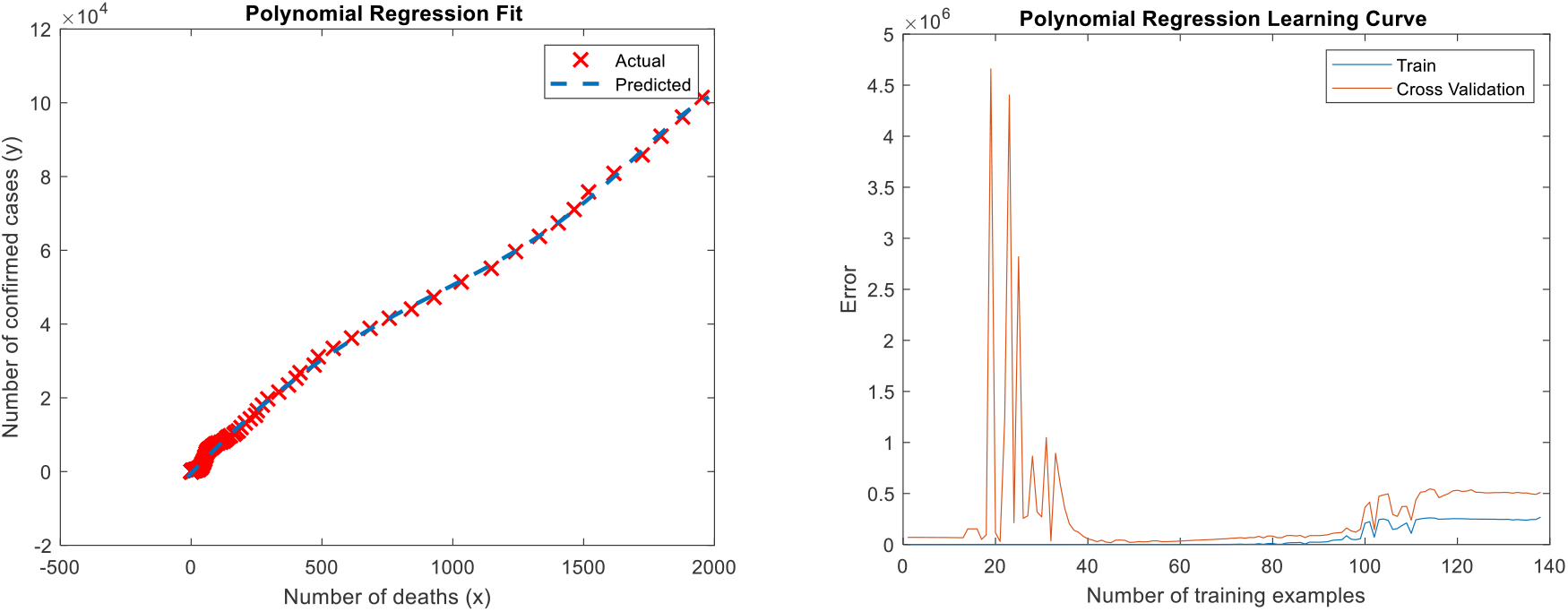
Polynomial Regression fit for state of Karnataka and Polynomial Regression Learning Curve.

**Figure 7:**
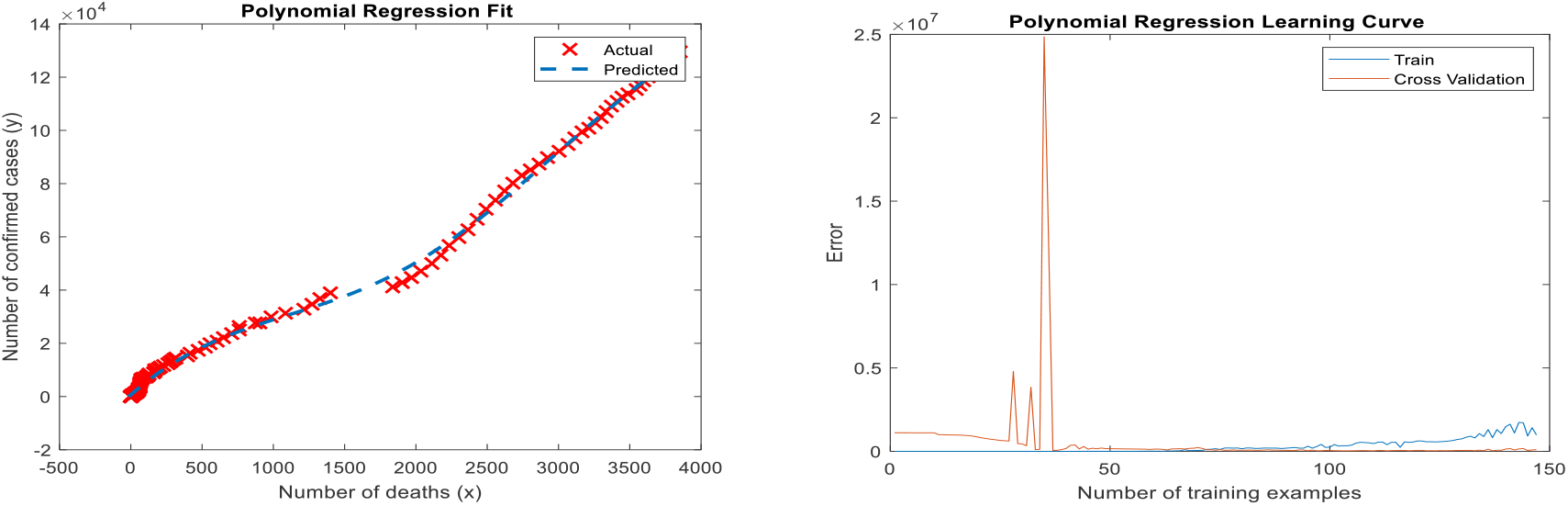
Polynomial Regression fit for state of Delhi and Polynomial learning Curve.

**Figure 8:**
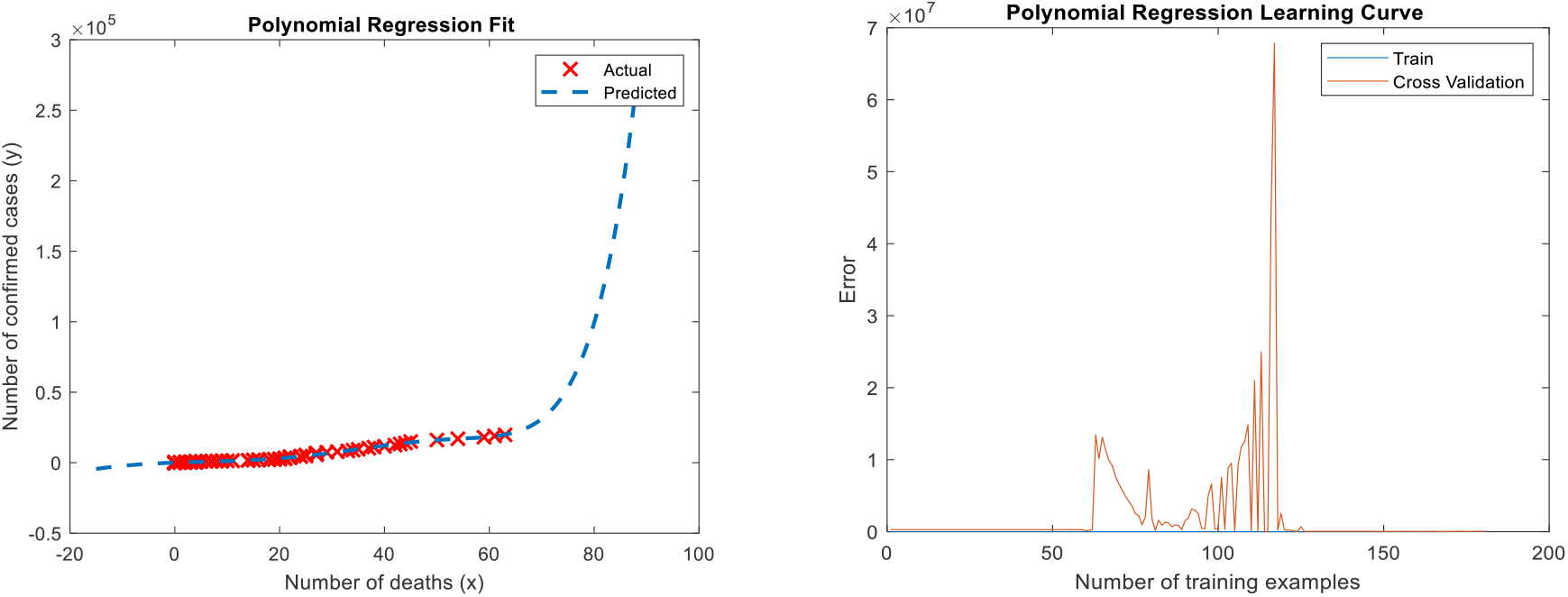
Polynomial Regression fit for state of Kerala and Polynomial learning Curve.

Polynomial regression is also done using kaggle dataset till 28^th^ July 2020.The predicted and actual results almost matches as shown in the graphs.

## 4. Data Analysis and Discussion

Regression analysis like Linear and polynomial is used to analyse the effect of covid 19 on different states of India.The results can vary depending upon the data set considered.In this paper, some of the states has been considered.This analysis can be extended to other states also.Also analysis can be done considering confirmed cases and recovered cases. Analysis shows that the number of confirmed cases and mortality rate is going to increase in the coming months as community spread has started. Maharashtra has the highest with 441228 confirmed cases and 15576 deaths.Andhra Pradesh has 158764 cases with 1474 deaths.Madhya Pradesh has 33535 confirmed cases and 886 deaths.Uttar Pradesh has 92921 confirmed cases and 1730 deaths.West Bengal has recorded 75516 confirmed cases and 1678 deaths.Rajasthan has 43804 cases and 703 deaths as on 3^rd^ August 2020.Mizoram and Lakshadweep has zero mortality has on 3^rd^ August.Almost all states are affected by Covid 19 and all measurements has to be taken to reduce the spread and to give accessibility to health care irrespective of whether rich or poor.

## 5. Conclusion

In this paper, Covid 19 cases has been analysed using dataset from kaggle website.Linear regression and polynomial regression has been done using the available dataset.This analysis shows that the number of cases are going to increase In the coming days.With respect to mortality rate, Maharashtra is the worst affected.Delhi and Tamil Nadu are aslo badly affected with respect to confirmed cases and mortality rate.Kerala has less mortality rate as compared to its confirmed cases. This study can be further extended by considering recovery rate and confirmed cases.Also other than regression, other models can also be used for analysis.

## Data Availability

Data was directly downloaded from github

https://github.com/imdevskp/covid-19-india-data

